# Prevalence of Carpal Tunnel Syndrome and its associated factors among patients with musculoskeletal compliant at Dilchora Referral Hospitals in Dire Dawa administration, Eastern Ethiopia, 2022

**DOI:** 10.1101/2023.02.10.23285779

**Authors:** Tewodros Yesuf, Hailu Aragie, Yared Asmare

**Affiliations:** Biomedical department, College of Medicine and Health Sciences, University of Dire Dawa, Dire Dawa, Ethiopia; School of Medicine, College of Medicine and Health Sciences, University of Gondar, Gondar, Ethiopia

**Keywords:** Carpal Tunnel Syndrome, Hand Dimension, Wrist Dimension and prevalence

## Abstract

**Background:** Carpal tunnel syndrome (CTS) is a chronic focal mono-neuropathy caused by mechanical distortion of the nerve at the carpal tunnel. It is thought to affect between 4 and 5 % of people worldwide, 50 per1000 persons in developed countries, 12.1% in east Africa and 29.2% Ethiopia. The common risk factors include but not limited to, age, sex, inflammatory conditions, pregnancy, diabetes mellitus, and hypertension. Despite this, carpal tunnel syndrome prevalence and its associated factors among patients with musculoskeletal complaints are unaddressed.

**Objectives:** This study’s aim was to assess the prevalence of carpal tunnel syndrome and its associated factors among patients with musculoskeletal complaints in Dire Dawa, Eastern Ethiopia.

**Methodology:** An institution-based cross-sectional study was conducted from June 1 to 30, 2022 at Dilchora referral hospital. 265 study participants were selected using the systematic sampling technique. Interviewing techniques and physical examination of the hands and wrists were used to get the data. Epi Data version 3.1 was used to enter, clean up, and edit the data before exporting it for analysis to SPSS version 23.0 software. Bivariable and multivariable logistic regression were carried out with a 95% confidence interval to identify the association of independent and dependent variables. A P-value of 0.05 was considered statistically significant.

**Result:** A total of 260 respondents were included in this study. The prevalence of clinically proven carpal tunnel syndrome among study participants was 10.8%, with a 95 % CI of (6.99 to 14.6). A multivariable analysis found that being female (AOR: 3.26 (95% CI: 1.05, 10.08), being physically inactive (AOR: 6.32 (95% CI: 1.95, 20.52), diabetes mellitus (AOR: 4.23 (95% CI: 1.47, 11.97)), hypertension (AOR: 6.07 (95% CI: 1.70, 21.65)), hand ratio ≤2.1 (AOR: 7.31(95% CI: 1.80, 29.66)), and wrist ratio ≥ 0.72 (AOR: 5.94 (95% CI: 2.11, 16.72)) were statistically associated factors of carpal tunnel syndrome. But, BMI were not statistically associated with CTS.

**Conclusion:** The prevalence of carpal tunnel syndrome among patients with musculoskeletal compliant was 10.8%. Several risk factors for CTS have been identified.

## Introduction

Musculoskeletal disorder (MSD) is an injury of the muscles, tendons, ligaments, nerves, joints, cartilage, bones, or blood vessels in the arms, legs, head, neck, or back that is caused or aggravated by certain medical conditions and tasks such as lifting, pushing, pulling, repetitive task, awkward posture. It is the most common cause of physical disability and severe long-term pain. Population surveys estimated that for a one-month period of recall, up to 50% of people in the general population experience musculoskeletal pain at one or more anatomical sites. Carpal tunnel syndrome is considered as one of the six common cause musculoskeletal disorder(1).

The carpal tunnel (CT) is found at the base of the hand. The transverse carpal ligament is a thick fibrous roof, and the eight carpal bones form part of its border (TCL). The tunnel allows for the passage of the thumb’s flexor pollicis longus (FPL) tendon, eight digital flexor tendons (two for each of the medial four fingers), their flexor synovial sheaths, and the median nerve (MN). Because of this, CT is relatively densely packed, and any circumstance that could increase the volume of the structures within it could result in MN compression. This may then result in the nerve’s ischemia, which manifests as pain and paresthesia(4).

Carpal tunnel syndrome (CTS) is the most common and well-known upper limb entrapment neuropathy, accounting for 90% of all entrapment neuropathies. It is characterized as a chronic focal compressive mono-neuropathy or radiculopathy caused by mechanical distortion of the nerve at the site where it passes through a narrow, non-flexible anatomical structure, which results in an ischemic response to the median nerve due to repetitive motion and high force. It frequently results in prolonged impairment that necessitates surgical intervention (5).

All signs of carpal tunnel syndrome include pain in the hand, unpleasant tingling, pain or numbness in the distal distribution of the median nerve (thumb, index, middle finger, and radial side of the ring finger), and a decline in grip strength and function of the affected hand are all signs of carpal tunnel syndrome(6,7). The symptoms are frequently worse at night, and daytime tasks involving wrist flexion are associated with clumsiness. Patients frequently speak of the “flick sign,” a phenomena in which shaking or flicking their wrists alleviates discomfort (8).

The diagnosis of CTS is made based on the presence of specific symptoms (hand paraesthesia, weakness and pain in the distribution of the median nerve); as well as positive results from a variety of provocative stress tests, including the Tinel’s, Phalen’s, and carpal tunnel compression tests; and confirmatory electrophysiology. The electrophysiological diagnostic criteria used were a distal motor latency (DML) of more than 4.0 milliseconds and/or a distal sensory latency of more than 3.5 milliseconds at the wrist level. Asymmetry in conduction times between both hands of greater than 1.0 milliseconds for motor conduction or 0.5 milliseconds for sensory conduction was also considered abnormal(9).

The question of whether a syndrome that is primarily responsible for the onset of CTS and its prevention is one that is occupationally caused or personally attributable has been the subject of intense discussion over the past few decades. In many instances, the cause of CTS is unknown, but several risk factors have been linked to its onset and exacerbation. Risk factors related to the environment are the most important. Long-term wrist flexion or extension postures, repetitive flexor muscle activity, vibration exposure, and any disease that congests or limits the capacity of the carpal tunnel might cause symptoms to appear. Among the most frequent causes of CTS are disorders such as misaligned Colles’ fracture, edema from infection or trauma, posttraumatic arthritis, or tumorous conditions including ganglion, lipoma, or xanthoma. Physical inactivity is a personal risk factor for CTS, and the syndrome can occasionally be linked to systemic diseases like obesity, diabetes, thyroid dysfunction, amyloidosis, and Raynaud’s syndrome(10–18).

A number of interrelated pathophysiologic mechanisms, such as increased carpal tunnel pressure, injury to the median nerve’s microcirculation, and changes to the connective tissue of the median nerve, as well as inflammation and hypertrophy of the synovial tissue, can explain the occurrence of carpal tunnel syndrome (CTS) (19).

Using univariate and multivariate analysis, the morphometric examination of the hand, wrist, and other body parameters has been examined to assess the likelihood of susceptibility to carpal tunnel syndrome (CTS). Positive correlations between several hand and body measurements have thus been repeatedly shown. Idiopathic carpal tunnel syndrome (ICTS) is more likely to occur in hands that are larger, thicker, and have squarer wrists, or “coarse” hands)(6,7,19–22).

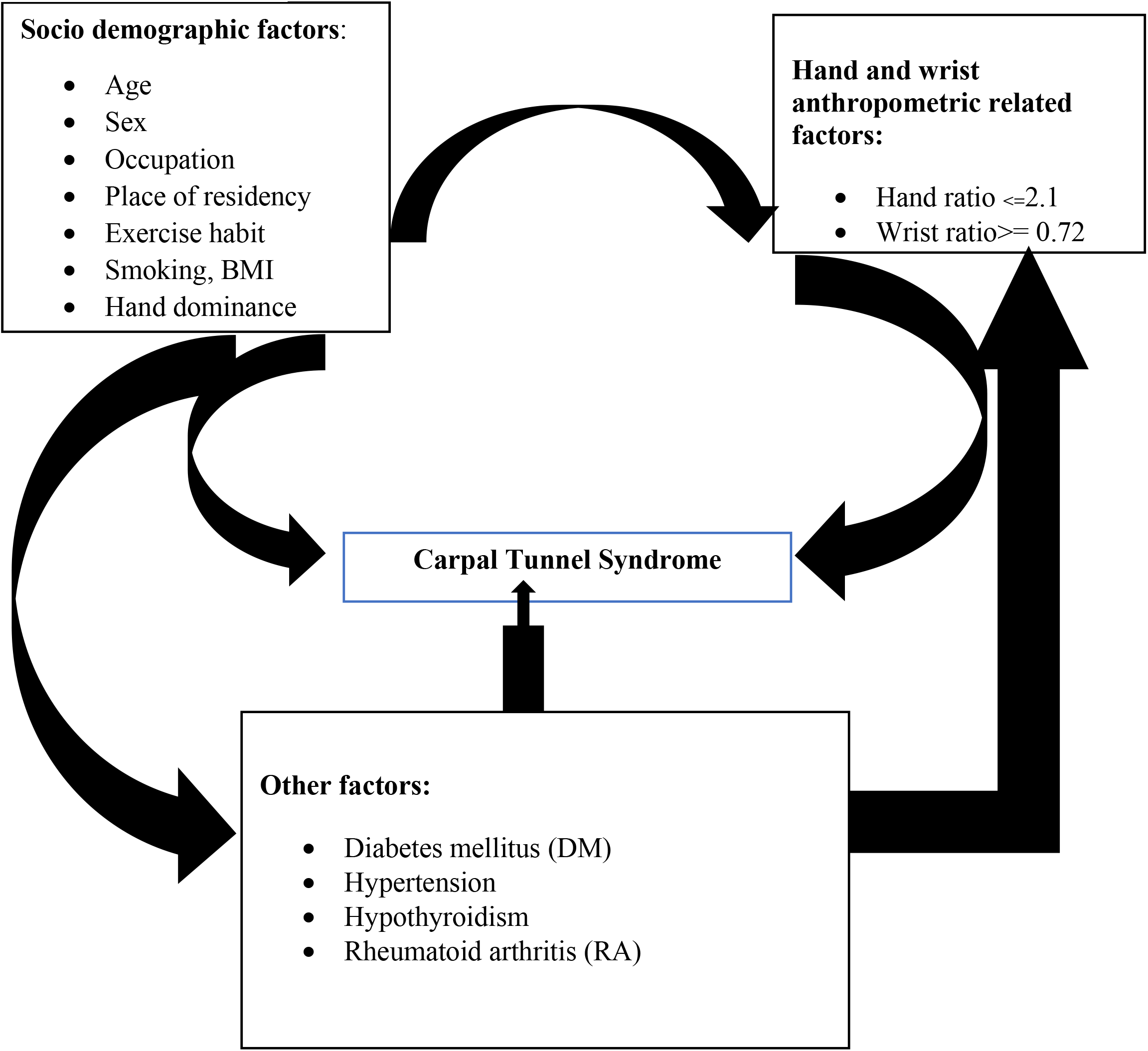
**Source**: constructed by the investigator from the literatures included in the study Conceptual framework on factors associated with carpal tunnel syndrome

## Objectives of the study

### General objective

To assess the prevalence of carpal tunnel syndrome and its associated factors among patients with musculoskeletal complaints attending medical and orthopedic outpatient department at Dill Chora referral hospital in Dire Dawa, Eastern Ethiopia, June 2022.

### Specific Objectives

1. To determine the prevalence of carpal tunnel syndrome among patients with musculoskeletal compliant attending the medical and orthopedic outpatient department.
2. To identify factors associated with carpal tunnel syndrome among patients with musculoskeletal compliant attending the medical and orthopedic outpatient department.

## Methods and materials

### Study design

An Institution-based cross-sectional study design was employed.

### Eligibility criteria

#### Inclusion criteria

- Patients who have musculoskeletal complaint and who are willing to participate the study.

#### Exclusion criteria

- Patients with congenital shoulder, elbow, and hand musculoskeletal abnormalities, recent fractures or injuries, and surgeries to the shoulder, elbow, and hand
- Patient with cognitive impairment that interfere communication.

### Sample Size Determination

The sample size for the first objective was determined by the formula for single population proportions, using the assumption of a 5% level of significance, P = Population proportion 50%, since no data related to the prevalence of CTS among patients with musculoskeletal complaints was available, marginal error of 5 %, and a 10% nonresponse rate. Considering the last three months of outpatient flow on average, a total of 1,932 clients were diagnosed with diseases of the musculoskeletal system (based on HMIS disease classification). So, 644 patients with musculoskeletal complaints were assumed to be available during one month of data collection. So, the sample was taken from a relatively small population (N = 644). The required sample size was obtained by the following calculation:

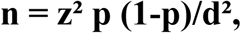

Where, n= sample size, p= prevalence, d= margin of error

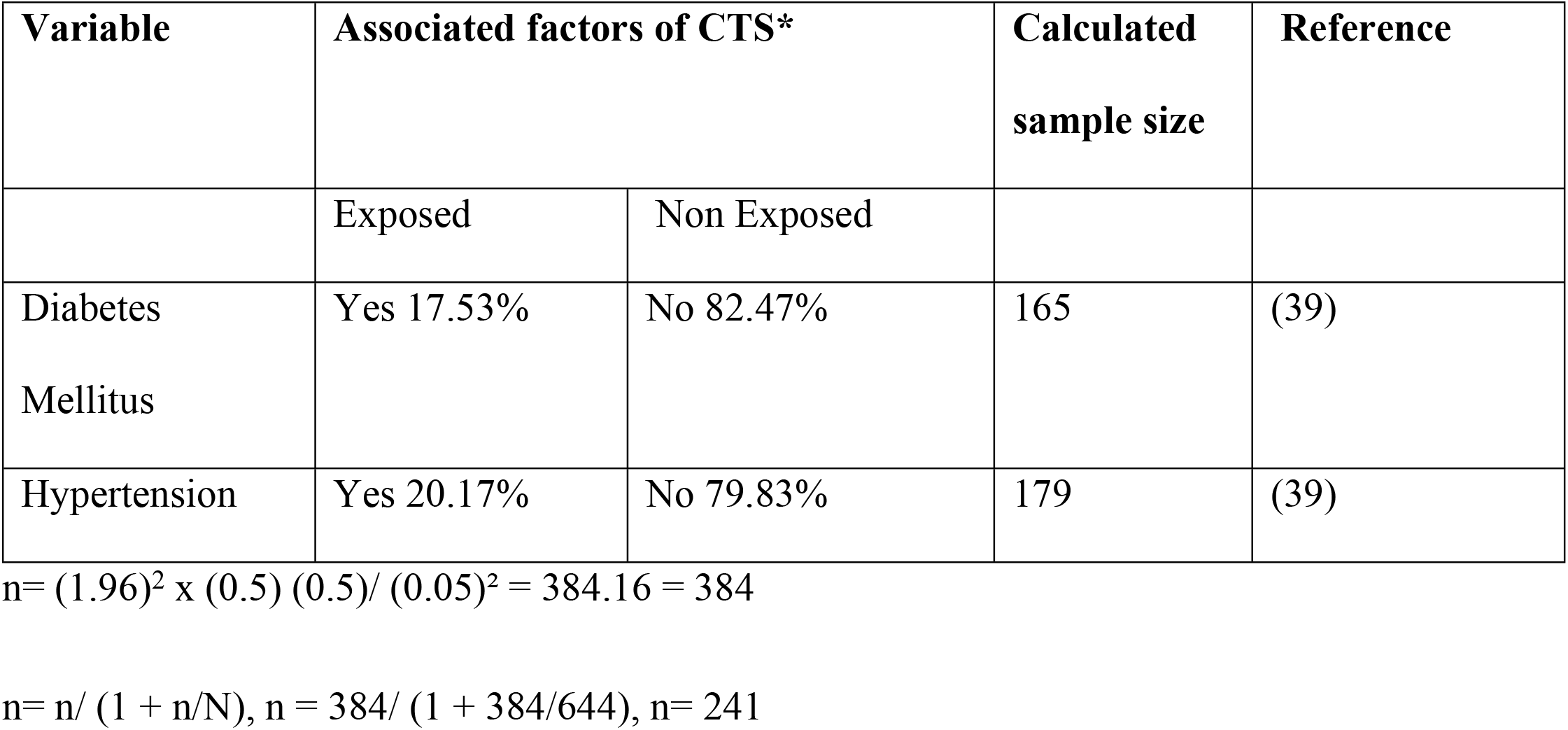

Sample Size Determination for the Second Objective

Double population proportion formula was used to determine the sample size for the factors associated with carpal tunnel syndrome. Sample size was calculated for some of the associated factors obtained from different literatures by using the Stat-calc of Epi Info statistical software version 7 with the following assumptions:

- Confidence level = 95%
- Power = 80%
- The ratio of unexposed to exposed almost equivalent to 1

Generally, sample sizes was calculated for the first and the second objectives and the largest sample size was 241 from the first objective and adding 10% for the non-response rate, the final sample size is **265**.

### Data collection tools and techniques

A face-to-face interviewer administered questionnaire was developed from relevant literature in the context of the study area. The questionnaire addressed (1) sociodemographic characteristics: age, gender, place of residency, exercise habits, smoking, and dominant hand; and (2) information related to previous history of medical illness, including: the presence of diabetes mellitus (DM), hypertension (HTN), hypothyroidism, and rheumatoid arthritis (RA). The data was collected by two data collectors and supervised by the principal investigator. The tools for the clinical diagnosis of CTS were performed by using a standard diagnostic questionnaire used in the previous study; a score of 5 or more is recommended for use of the test as a diagnostic tool to replace nerve conduction studies, which is the gold standard test(42).

In addition to the standard diagnostic questionnaire, two important clinical tests were performed to increase the case detection rate. (1) Phalen’s wrist flexion test: passively flexing the wrist for 60 seconds produces pain and paraesthesia in the median nerve distribution and implies a positive result. The test has a sensitivity of 68% and a specificity of 73%. (2) Carpal tunnel compression test: applying firm pressure directly over the carpal tunnel, usually with the thumbs, for up to 30 seconds to produce symptoms and imply a positive result. This test has a sensitivity of up to 89% and a specificity of 96 %(18, 43).

### Data quality control

The questionnaire was pretested before the actual data collection period on 5% of the sample size in Hiwot Fana Specialized University Hospital (HFSUH) to ensure its consistency and validity. The questionnaire was amended after the pretest. Two days of training were given to the data collectors on the data collection tool and the data collection procedure by the principal investigators. The completeness of each questionnaire was checked by the investigator on a daily basis. Double data entries were done by two data clerks, and the consistency of the entered data was cross-checked by comparing the two separately entered data on EpiData.

### 3.11. Data analysis and interpretation

The collected data was checked for its completeness, coded and entered into Epi-Data version 3.1 statistical packages than were exported to SPSS version 23 for further analysis. Descriptive statistics of different variables were presented by frequency and percentage using tables, bar graphs and pie charts. For descriptive numerical variables mean and standard deviation was determined. Model fitness was cheeked by Hosmer-Lemeshow goodness of fit. Bivariable logistic regression analyses were applied to determine independent variables associated with CTS. Independent variables that with p-value <0.25 and/or variables shows an association with CTS in previous studies were illegible to multivariable logistic regression to control for the effect of confounders. Finally, significantly associated independent variables at p-values < 0.05 was identified based on the adjusted odds ratio (AOR), with 95% CI.

### Ethical considerations

The study was carried out after getting approval from the Institution Review Board University of Gondar. An Official letter was written to the Dilchora referral hospital by the University of Gondar College of medicine and health sciences and delivered to the facility directors to get permission. The purpose and objective of the study were elaborated to each informant and verbal consent was obtained from each study participants. The participants were also be informed that participation was voluntarily and they well aware of their rights that they could stop or leave participation at any time they felt any inconvenience or discomfort. To keep the confidentiality of any information provided by study subjects, the data collection procedure was anonymous.

## Result

### Socio-demographic Characteristics

A total of 260 respondents participated in the study. Of these 156 (60 %) women and 115(44.2%) are house wife. The mean age of the respondents was 52 (SD: 12.4) and majority of the participants, 206 (79.1%) were urban dwellers. (**Table 1**)

**Table 1:**
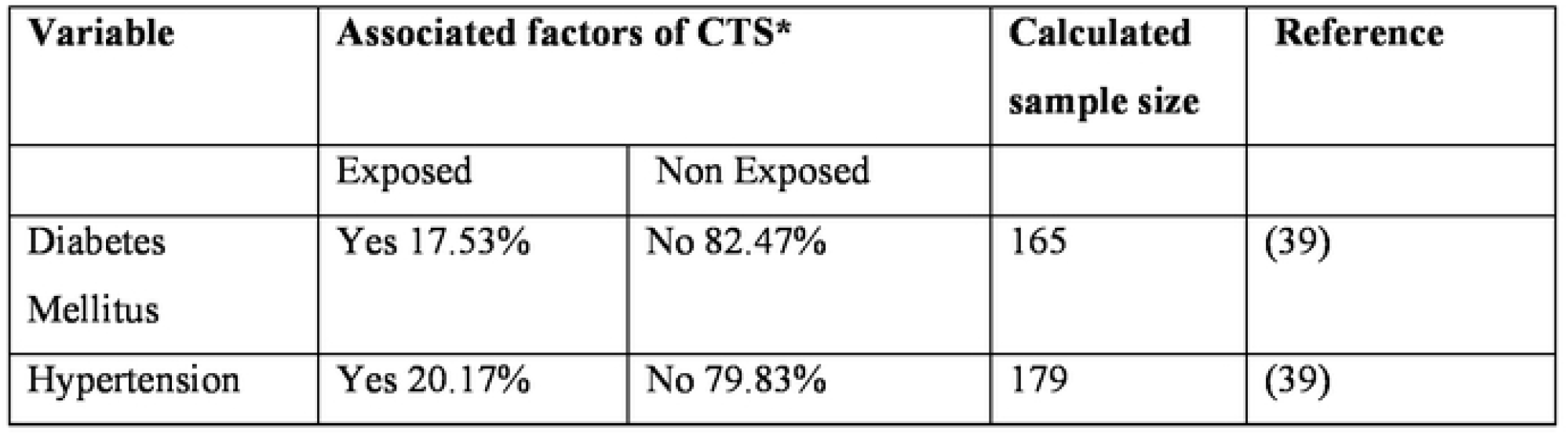
Sample size calculation for factors associated with carpal tunnel syndrome

### Health characteristics of respondents

One hundred twenty-five (48.1%) of the participants had history of past illness. Of these 78 (30 %) had diabetes mellitus and 36 (13.8%) had hypertension. **(Figure 1)**

**Figure 1:**
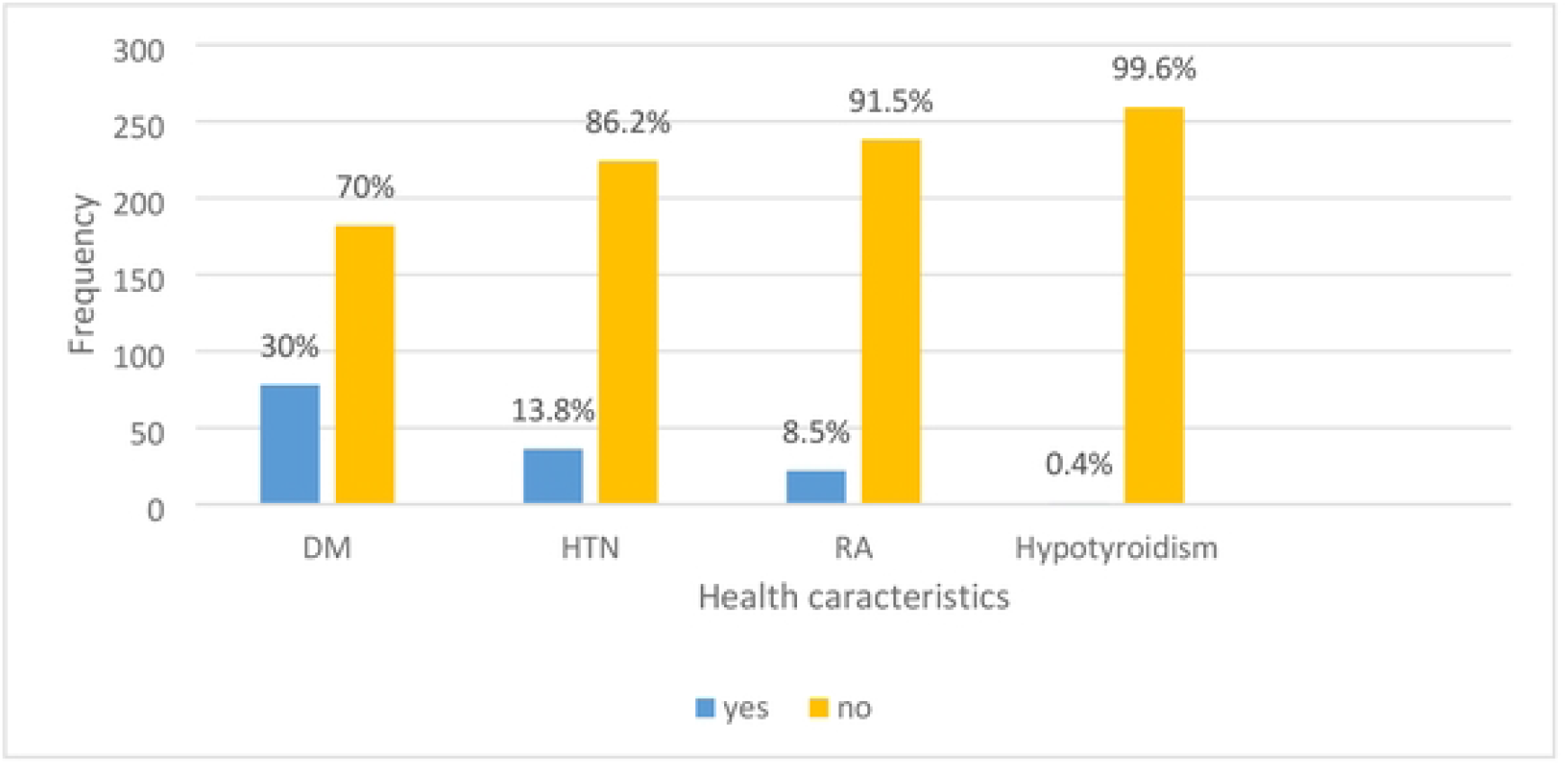
Health traits of study participants attending Dilchora referral hospital, Eastern Ethiopia, 2022.

Regarding current musculoskeletal complaints of the participants, 119 (45.8%) experienced neck pain. Of these, 12(10.1%) had clinically confirmed CTS and 56 (21.5%) of the participants complained of wrist and hand pain. Of these, 16(28.6%) had clinically confirmed CTS. **(Table 2)**

**Table 2:**
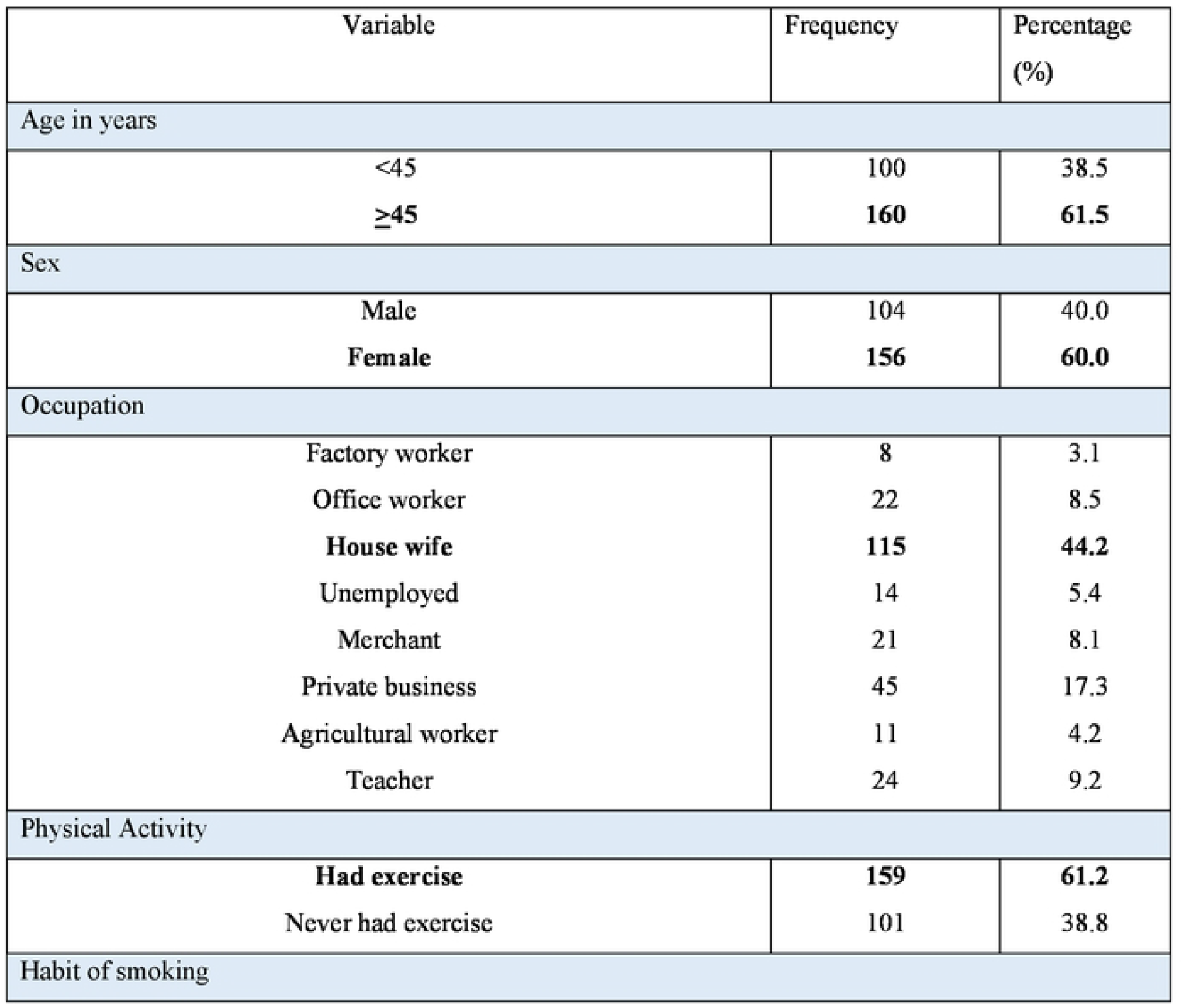
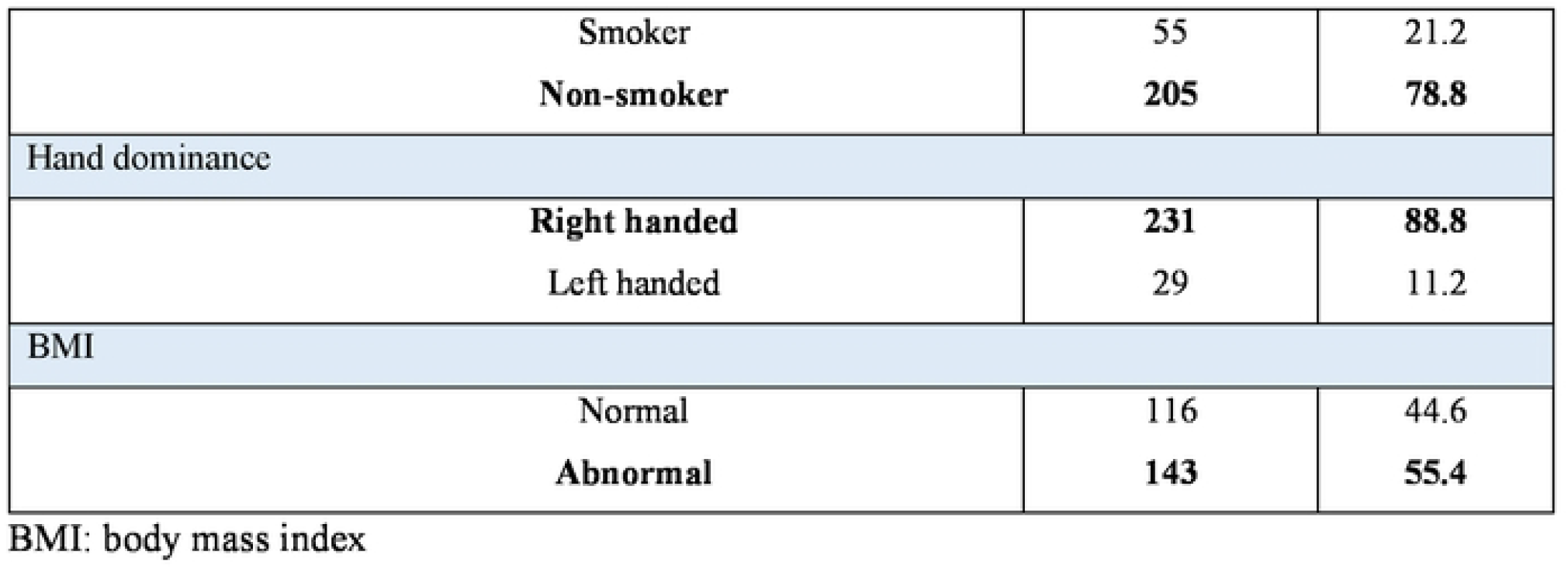
Socio-Demographic Characteristics of Study Subjects for Prevalence of Carpal Tunnel Syndrome and its associated factors among patients with a musculoskeletal complaint in Dilchora referral Hospital, Eastern Ethiopia, 2022. (n=260)

### Total score for participant response to clinical diagnostic criteria

Of the total study participants, 146 (56.1%) had symptoms of carpal tunnel syndrome. Of these 28 (10.8 %) patients had a score of more than or equal to 5, which is the threshold for a clinical diagnosis of CTS. One hundred eighteen (45.4%) of the study subjects had clinically uncertain score to diagnose the presence of CTS. (**Table 3**)

**Table 3:**
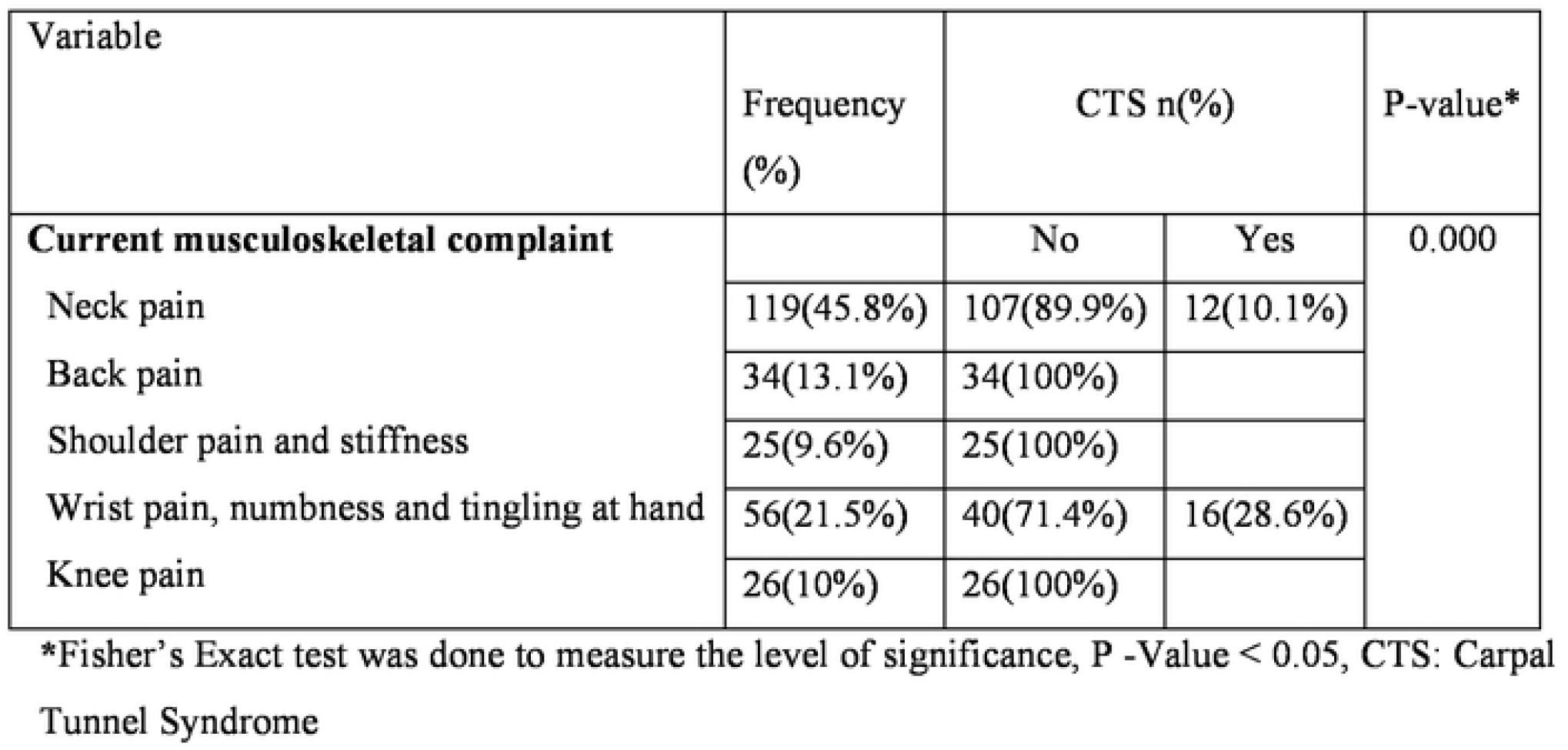
Musculoskeletal skeletal complaint of the participants attending Dilchora referral hospital, Eastern Ethiopia, 2022.

| Variable | Frequency (%) | CTS n(%) |  | P-value* |
| --- | --- | --- | --- | --- |
| **Current musculoskeletal complaint** |  | No | Yes | 0.000 |
| Neck pain | 119(45.8%) | 107(89.9%) | 12(10.1%) |  |
| Back pain | 34(13.1%) | 34(100%) |  |  |
| Shoulder pain and stiffness | 25(9.6%) | 25(100%) |  |  |
| Wrist pain, numbness and tingling at hand | 56(21.5%) | 40(71.4%) | 16(28.6%) |  |
| Knee pain | 26(10%) | 26(100%) |  |  |

\*Fisher's Exact test was done to measure the level of significance, P -Value < 0.05, CTS: Carpal Tunnel Syndrome

Regarding the clinical examination of the participants, 54 (20.8%) of the subjects experienced pain and paraesthesia in the median nerve distribution while passively flexing the wrist. Only 18 (33.3%) of them had CTS that is consistence with clinically proven CTS. **(Table 4)**

**Table 4:** The overall score in response to the clinical diagnostic criterion for prevalence of carpal tunnel syndrome and its associated factors among patients with musculoskeletal complaint in Dilchora referral hospital, Eastern Ethiopia, 2022.(n=260)

| Total score | Frequency | Percent (%) | Interpretation |
| --- | --- | --- | --- |
| A score of "0" | 114 | 43.8 | Wrist and hand symptom is not caused by |

|  |  |  | CTS |
| --- | --- | --- | --- |
| A score of “<3” | 114 | 43.8 | Unlikely to be indicative of Carpal Tunnel Syndrome |
| A score of “3-4” | 4 | 1.6 | Suggests Carpal Tunnel Syndrome is possible cause of symptoms |
| A score of “≥5” | 28 | 10.8 | Strongly suggestive of Carpal Tunnel Syndrome( **Define the presence of CTS**) |

### Measured anthropometric parameters of hand and wrist dimension

Mean (SD) wrist ratio and hand ratio for the participants were 0.60(SD: 0.113) and 2.40(SD: 0.218) respectively. **(Figure 2)**

**Figure 2:**
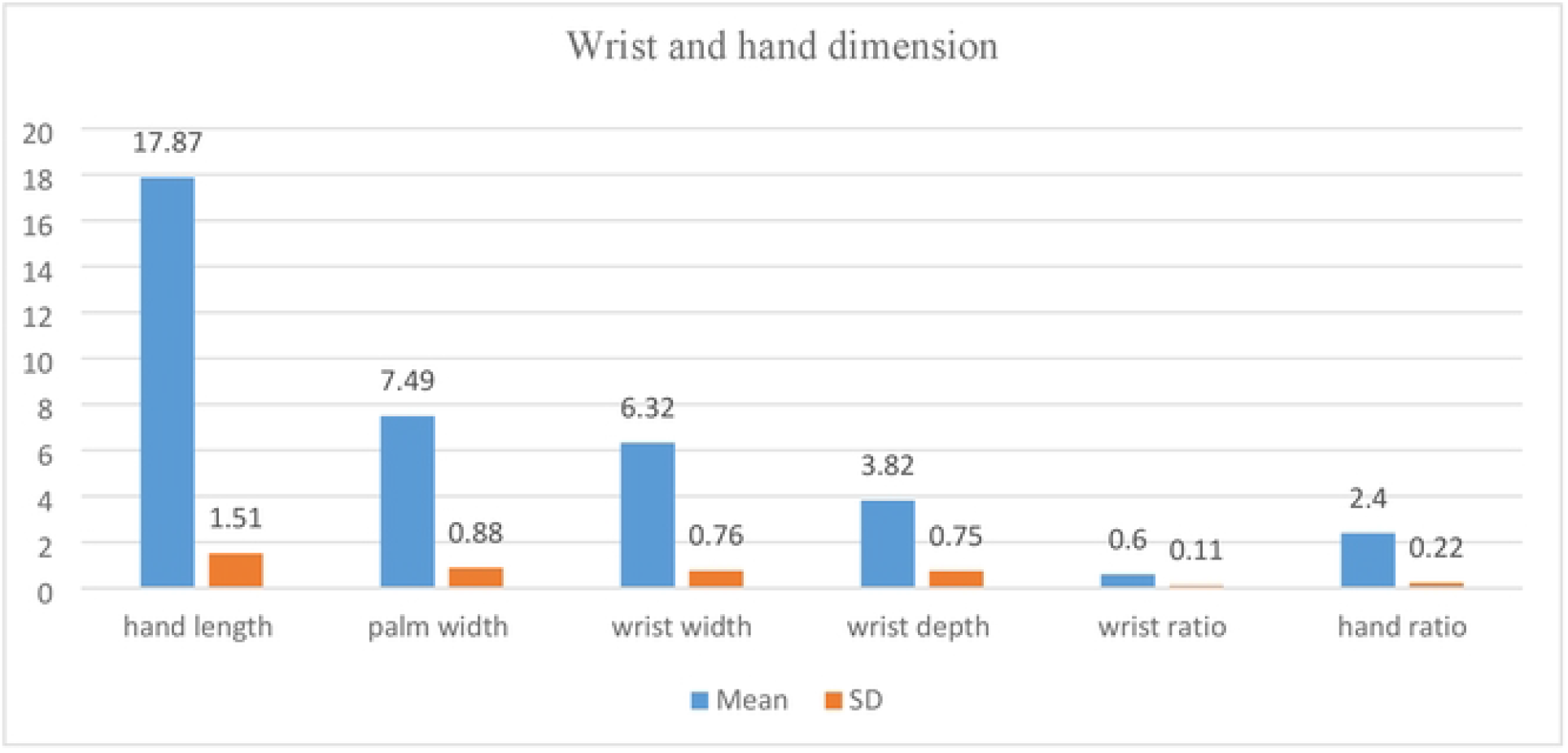
Mean and standard deviation of wrist and hand dimension of the study participants attending Dilchora referral hospital, Eastern Ethiopia, 2022.

Regarding the hand and wrist index 56 (21.5%) of the participants were found to have square wrists, which are those with a wrist ratio greater than or equal to 0.72. Of a total of the participants 147(56.5%) found to have shorter and wider hands. **(Figure 3)**

**Figure 3:**
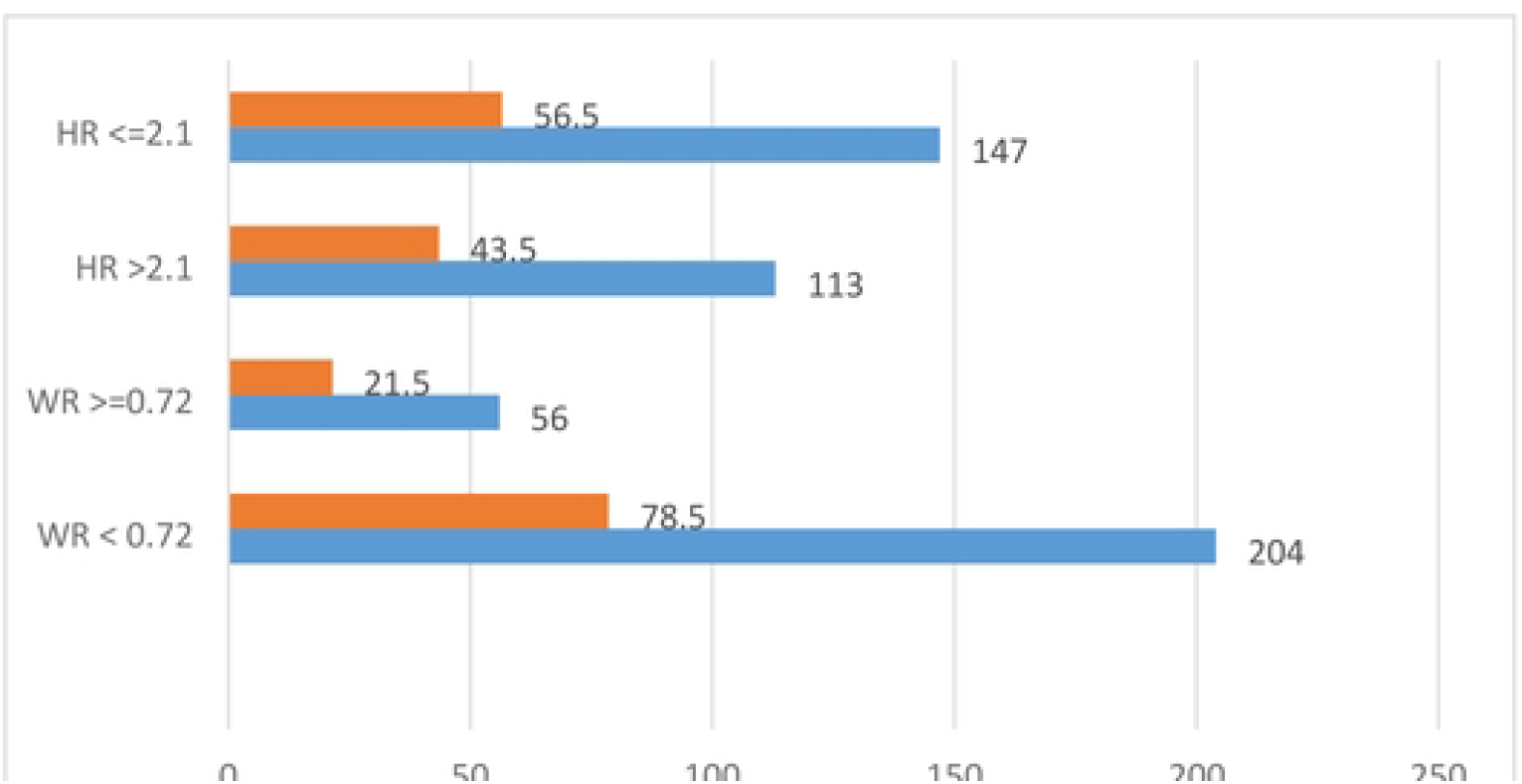
Distribution of hand and wrist ratio of study participants attending Dilchora referral hospital, Eastern Ethiopia, 2022.

### Prevalence/Magnitude of clinically confirmed CTS

The prevalence of clinically proven carpal tunnel syndrome among study participants was 28 (10.8%) with a 95 % CI: (6.99, 14.6%). Females had the highest frequency, at 22 (14.1%). Age 45 years and older had a higher prevalence of CTS, 19(11.9%). (**Table 5)**

**Table 5:** The proportion of participants who had medical examination to ascertain the prevalence of carpal tunnel syndrome and its associated factors among patients with musculoskeletal complaint in Dilchora referral hospital, Eastern Ethiopia, 2022. (n=260)

| Type of test | Test Result | Frequency (%) | CTS Status |  |
| --- | --- | --- | --- | --- |
|  |  |  | Yes | No |
| **Phalen’s wrist flexion** | + | 54(20.8) | 18(33.3%) | 36(66.7%) |
|  | - | 206(79.2) | 10(4.9%) | 196(95.1%) |
| **Carpal Tunnel Compression** | + | 56(21.5) | 19(33.9%) | 37(66.1%) |
|  | - | 204(78.5) | 9(4.4%) | 195(95.6%) |

Chi square test was done to measure the level of significance, +: Positive, - : Negative, CTS: carpal tunnel syndrome,

### Factors associated with carpal tunnel syndrome

Carpal tunnel syndrome was compared with sex, diabetes mellitus, hypertension, wrist ratio greater than or equal to 0.72, hand ratio less than or equal to 2.1, body mass index greater than or equal to 25 kg/m2, and participants’ exercise habit in a bivariate logistic regression analysis (p0.05). Before controlling for confounders, the analysis’s findings show that there was a significant association. **(Table 6)**

**Table 6:** The prevalence of CTS by participant’s age, gender and past medical illness (n=260)

| Variable | CTS |  | P-Value* |
| --- | --- | --- | --- |
|  | No, n (%) | Yes, n (%) |  |
| Sex |  |  |  |
| Male | 98(94.2%) | 6(5.8%) | 0.034 |
| Female | 134(85.9%) | 22(14.1%) |  |
| Age |  |  |  |
| <45 | 91(91.0%) | 9(9.0%) | 0.467 |
| <u>=</u>>45 | 141(88.1%) | 19(11.9%) |  |
| Past medical condition |  |  |  |
| DM |  |  | 0.02 |
| Yes | 66(80.5%) | 16(19.5%)) |  |
| No | 166(93.3%)) | 12(6.7%) |  |
| HTN |  |  | 0.02 |
| Yes | 103(83.1%)) | 21(16.9%) |  |
| No | 129(94.9%) | 7(5.1%) |  |

\* Chi-square test, P-Value <0.05, CTS: carpal tunnel syndrome, DM: diabetes mellitus, HTN: hypertension

**Table-7.** Bivariate relationship between selected variables with carpal tunnel syndrome at Dilchora referral Hospital, Dire Dawa, Eastern Ethiopia, 2022. (n=260)

| Variable | Carpal tunnel syndrome |  | COR(95%CI) | p-value |
| --- | --- | --- | --- | --- |
|  | No, n | Yes, n |  |  |
| Sex |  |  |  |  |
| Female | 134(85.9%) | 22(14.1%) | 2.682(1.048,6.862) | 0.040 |
| Male | 98(94.2%) | 6(5.8%) | 1.00 |  |
| Exercise habit |  |  |  |  |
| Never had exercise | 84(83.2%) | 17(16.8%) | 2.723(1.218,6.086) | 0.015 |
| Had exercise | 148(93.1%) | 11(6.9%) | 1.00 |  |
| Diabetes mellitus |  |  |  |  |
| Yes | 66(80.5%) | 16(19.5%) | 3.354(1.505,7.471) | 0.003 |
| No | 166(93.3%) | 12(6.7%) | 1.00 |  |
| Hypertension |  |  |  |  |
| Yes | 103(83.1%) | 21(16.9%) | 3.757(1.537,9.184) | 0.04 |
| No | 129(94.9%) | 7(5.1%) | 1.00 |  |
| Body mass index |  |  |  |  |
| BMI* |  |  |  |  |
| >25kg/m<sup>2</sup> | 95(83.3%) | 19(16.7%) | 3.044(1.321,7.018) | 0.009 |
| ≤24.99 | 137(93.8%) | 9(6.2%) | 1.00 |  |
| Hand and wrist anthropometry |  |  |  |  |
| HR* |  |  |  |  |
| ≤2.1 | 122(83%) | 25(17%) | 7.514(2.20,25.57) | 0.01 |
| >2.1 | 210(97.3%) | 3(2.7%) | 1.00 |  |
| WR* |  |  |  |  |
| ≥0.72 | 42(75%) | 14(25%) | 4.524(2.07,10.196) | 0.000 |
| <0.72 | 190(93.1%) | 14(6.9%) | 1.00 |  |

**WR\*: wrist ratio, HR\*: hand ratio, BMI\*: body mass index**

**Table 8:**
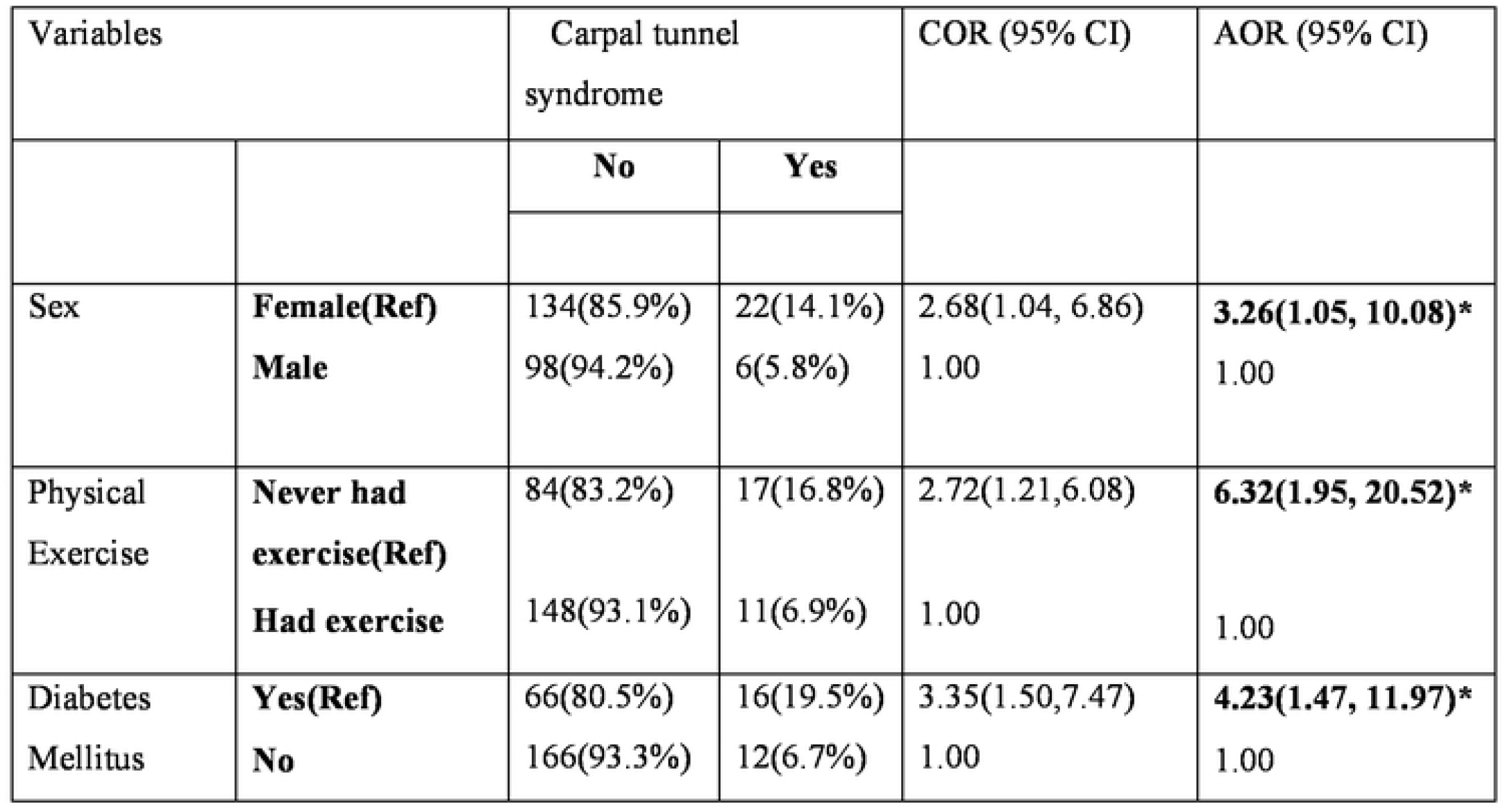
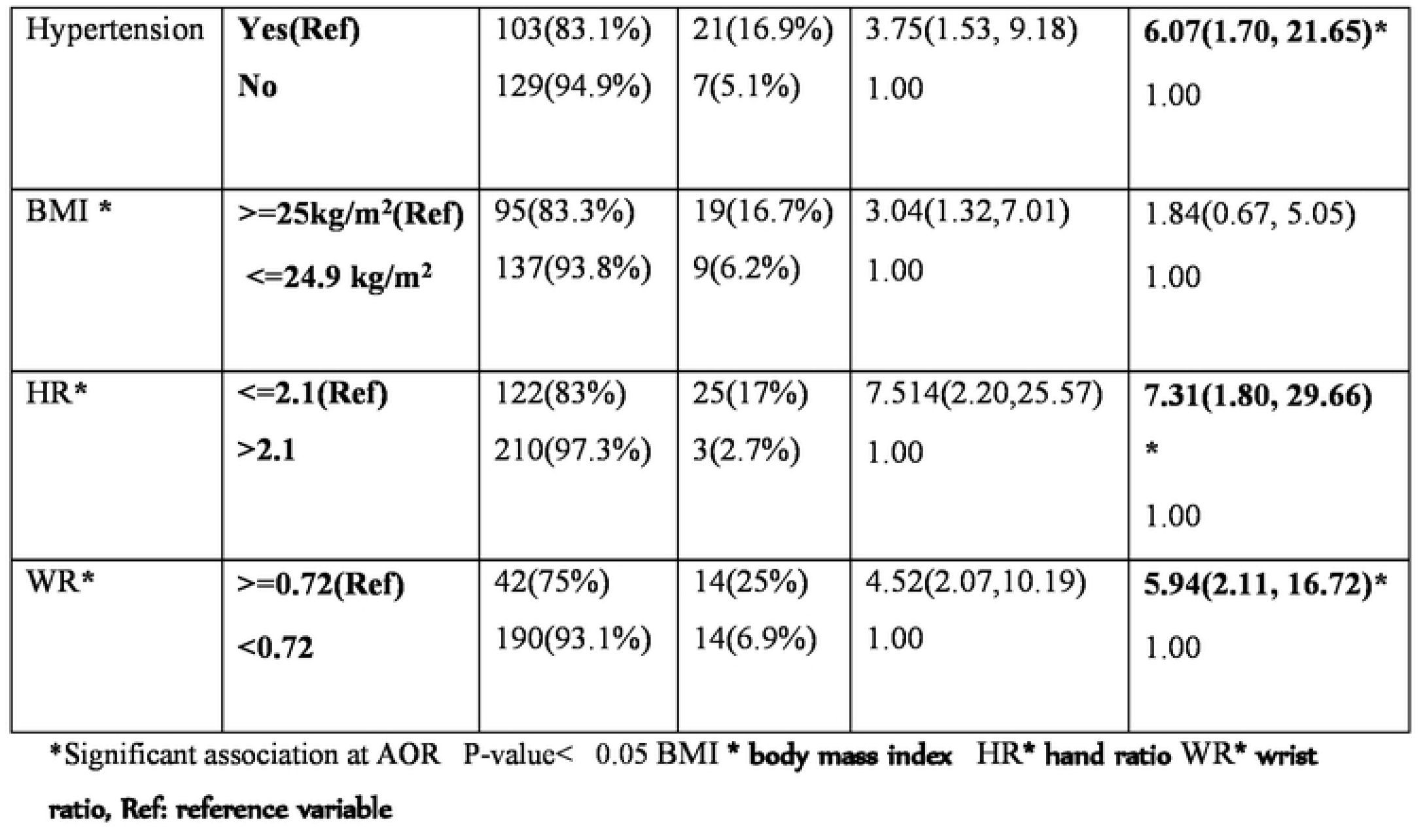
Multiple logistic regression of carpal tunnel syndrome with predictor variables in Dilchora referral hospital, Dire Dawa, Eastern Ethiopia, 2022. (n=260)

## Discussion

The results of this study indicate that the prevalence of clinically confirmed carpal tunnel syndrome among patients with musculoskeletal complaints was 28 (10.8%), with a 95 % CI: (6.99 to 14.6). Female gender, being physically inactive or never having exercised, the presence of diabetes mellitus, the presence of hypertension, hand ratio ≤ 2.1 and wrist ratio ≥ 0.72 were found to be risk factors for carpal tunnel syndrome.

The results of this study are consistent with findings from studies on diabetes mellitus patients carried out in Iran and Saudi Arabia. There were 8.56% and 6.7% prevalence rates, respectively(44,45). The outcome of this study is also consistent with a study among office employees in China, where the prevalence of clinically confirmed CTS was 9.6%(24).

In contrast, the result of the present study is lower than the findings of the studies conducted among diabetic patients in the United Kingdom (20%), India (19.8%), Bangladesh (26%), and Ethiopia (29.2%)(28,37,46,47). Similarly, the prevalence of CTS in this study was much lower than the findings of the studies conducted in Saudi Arabia (81.7%) and Korea (71.2%)(9,48). This variation in prevalence may be attributed to the chosen criteria to define the cases as CTS. In the previous studies, a more sensitive electro-diagnostic criterion that compares differences between distal median and ulnar distal sensory latencies was adopted, which can increase the case detection rate. Additionally, all the previous studies used diabetic patients as the study population, which is one of the common risk factors for developing carpal tunnel syndrome and the specific relationship of CTS to diabetes is thought to be due to median nerve entrapment caused by diabetes induced connective tissue changes.

The 10.8% prevalence of carpal tunnel syndrome in the present study is higher as compared with studies conducted in Arbaminch general hospital (3.1%), Tikur anbesa specialized hospital (1%) and Jordan (5.5%) (26,49,50). This difference in prevalence might be due to the case definitions based on symptoms alone in this specific study, leading to relatively higher estimates of disease prevalence compared to definitions requiring both symptoms and electro-diagnostic study confirmation.

In accordance with the risk factors for developing carpal tunnel syndrome among the participants in the current study who experienced musculoskeletal complaints, women were 3.26 times more likely than men to do develop carpal tunnel syndrome. Likewise, a studies in South Korea and China found that women are 2 to 4 times more likely than men to develop CTS (24,51). Similar findings from an Iranian study indicate that women are 9.05 times more likely than men to develop carpal tunnel syndrome (32). This might be due to women having smaller carpal tunnel areas and volumes than men. Naturally smaller wrists of women may predispose them to the development of CTS(52). Moreover, it might be related to hormone level fluctuations. Estrogen receptors are located in the nervous system and their relevant actions are neuron proliferation and regeneration. As a result, estrogen levels are lower in premenopausal women and can affect the medial nerve (7).

According to the results of the current study, those who have never exercised are 6.23 times more likely to develop carpal tunnel syndrome than people who engage in physical exercise. Such a finding is consistent with the results of studies conducted in Saudi Arabia that showed the odds of having CTS was less than 1 among participants performing physical exercise occasionally (OR=0.11) (27). The result of this study is also supported by the findings of a case-control study conducted in the United States reporting that the odds of having CTS were significantly decreased in participants engaging in physical activity(53). This might be due to performing exercises can strengthen the muscles and improving blood flow to the wrist and hand.

Study participants in the current study who had diabetes as a health-related predictor had 4.23 times more likely to develop carpal tunnel syndrome as compared to those with no history of diabetes. This result is supported by previous studies, which reported that patients with DM were three times more vulnerable to CTS compared to the healthy population(51,54). Similarly, A nationwide population-based cohort study conducted in Taiwan revealed that patients with DM had the highest risk for diabetic hand syndromes (DHS), including CTS(55). This might be due to an increased serum glucose level resulting in attachment of the glucose to the proteins in the tendons located in the carpal tunnel. This will, in turn, cause the tendons to become inflamed and result in focal adherence. Moreover, there is also demyelination of the median nerve and axonal degeneration, macrophage attraction and activation, release of inflammatory cytokines, nitric oxide, and the development of “chemical neuritis,” which are all consequences of longstanding hyperglycemia(12).

The finding of the present study revealed that, those who had hypertension as health-related predictor had 6.07 times more likely to develop carpal tunnel syndrome as compared to those with no history of hypertension. This finding also supported by a study done in Boston indicated that having hypertension increase the risk of developing CTS by 1.88 times compared with non-hypertensive individuals(56). This might be due to a long-standing high blood pressure resulting in a breakdown in the blood-nerve-barrier formed by the inner cells of the perineurium and the endothelial cells of endoneurial capillaries that accompany the median nerve, increasing the permeability and causing an accumulation of proteins and inflammatory cells, contributing to increased endoneurial fluid and the development of edema(7).

The result of the present study also found that there was no significant association between body mass index (overweight and obese participants) and carpal tunnel syndrome. This finding is inconsistent with the report of a study conducted in Michigan Medical Center, United States, which indicated that, the odds ratio of obese (BMI > 29 kg/m^2^) compared to slender (BMI <20 kg/m^2^) individuals was 8.2 more likely to develop CTS and overweight (BMI 25-29 kg/m^2^) and obese individuals were 2.9 times more likely to develop CTS as compared to slender and normal (BMI 20-25 kg/m^2^) individuals(58). These discrepancies may be a result of the criteria that were used to define CTS patients in the current study, which may have resulted in these relationships among subjects with normal BMI. Additionally, the study’s participants are over 45 years old, which puts them at risk for carpal ligament degeneration. Patients with a normal BMI might develop CTS as a result of this. In contrast to this, the finding of the present study is comparable with that of a study conducted in India that showed that BMI was not associated with CTS(47).

From this study, it was found that patients with a hand ratio of ≤2.1 had 7.31 times more likely to develop carpal tunnel syndrome. This finding is supported by an experimental study conducted in Austria as a result, the hand ratio was significantly higher in the control group (Hand Ratio: 2.20) as compared with the patients’ group (Hand Ratio: 2.0)(19). This might be due to the assumption that shorter and wider hands (hand ratio ≤ 2.1) need extra force during normal hand movement, which leads to increased pressure in the intracarpal area, leading to CTS development (32).

The current study found that patients with a wrist ratio greater than 0.72 were 5.94 times more likely to develop carpal tunnel syndrome. This finding is consistent with the result of a meta-analysis of 16 studies, which concluded that a wrist ratio above 0.72 was associated with a three-fold increased risk of CTS(59). Similarly, a study conducted in Turkey showed that patients with a wrist ratio higher than 0.69 were 8.2 times more likely to have CTS (41). The result of this study is also supported by a cohort study that confirms 99% of normal laborers with square wrists (Wrist Ratio > 0.75) develop CTS (60). Moreover, the finding of this study was supported by an experimental study conducted in Greece as a result, the wrist ratio was significantly correlated with the carpal tunnel ratio (CTR) (22). This might be due to the assumption that the more square the wrist shape, the greater volar extension or flexion is required for a given motion, and this leads to median nerve compression and the development of CTS(17).

## Conclusion and recommendations

### Conclusion

The prevalence of carpal tunnel syndrome among patients with musculoskeletal complaints was 10.8%.

Being female, being physically inactive or never had exercise, presence of diabetes mellitus, the presence of hypertension, hand ratio ≤ 2.1 and wrist ratio ≥ 0.72 are the common risk factors identified in this study. There were statistically significant (P-value <0.05) association with the outcome variable (CTS).

### Recommendations

It is necessary to do additional research by increasing the number of the participants.

The assessment of the wrist and hand dimensions should then be a crucial component of care for patients with musculoskeletal complaints who also have diabetes mellitus and high blood pressure.

## Data Availability

All relevant data are within the manuscript and its Supporting Information files.

## ANNEX

